# Benefit versus Risk of Endomyocardial Biopsy for Heart Transplant Patients in the Contemporary Era

**DOI:** 10.1101/2023.05.19.23290196

**Authors:** Vincenzo Cusi, Florin Vaida, Nicholas Wettersten, Nicholas Rodgers, Yuko Tada, Bryn Gerding, Barry Greenberg, Marcus Anthony Urey, Eric Adler, Paul J. Kim

## Abstract

**Background:** The reference standard of detecting acute rejection (AR) in adult heart transplant (HTx) patients is an endomyocardial biopsy (EMB). The majority of EMBs are performed in asymptomatic patients. However, the benefit of diagnosing and treating AR compared to the risk of EMB complications has not been compared in the contemporary era (2010-current).

**Methods:** The authors retrospectively analyzed 2,769 EMB obtained in 326 consecutive HTx patients between August 2019 and August 2022. Variables included surveillance versus for cause indication, recipient and donor characteristics, EMB procedural data and pathologic grades, treatment for AR, and clinical outcomes.

**Results:** The overall EMB complication rate was 1.6%. EMBs performed within 1 month after HTx compared to after 1 month from HTx showed significantly increased complications (OR = 12.74, p < 0.001). The treated AR rate was 14.2% in the for cause EMBs and 1.2% in the surveillance EMBs. We found the benefit/risk ratio was significantly lower in the surveillance compared to the for cause EMB group (OR = 0.05, p < 0.001). We also found the benefit to be lower than risk in surveillance EMBs.

**Conclusions:** The yield of surveillance EMBs has declined, while for cause EMBs continued to demonstrate a high benefit/risk ratio. The risk of EMB complications was highest within 1 month after HTx. Surveillance EMB protocols in the contemporary era may need to be re-evaluated.

## Introduction

Acute rejection (AR) has been historically associated with early death after heart transplantation (HTx). Due to the initially high morbidity and mortality of AR, endomyocardial biopsy (EMB) was developed to detect AR early in HTx patients.^1^ Although recent advancements in noninvasive blood-based biomarkers show promise in replacing surveillance EMBs,^2–4^ EMB continues to be used for surveillance of AR in asymptomatic patients at most institutions in the first year after HTx.

Previous studies have described various complications associated with EMBs that range from 1% to 5% in HTx patients.^5–9^ While EMB complication rates remain unchanged, the incidence of AR detected by EMBs has decreased from 54% to 5%.^10, 11^ Deaths due to AR have also decreased.^12^ This shift has been attributed to advances in post-HTx care, particularly improved immunosuppression regimens.

Because of the marked reduction in AR and also the concern for over immunosuppression,^13^ the role for surveillance EMB in HTx patients is being re-evaluated.^14, 15^ However, a direct comparison of the benefit and risk of EMBs in both surveillance and for cause EMBs has not been performed in the contemporary era (2010-current).

In the present single-center study, we compared the rate of treated AR versus EMB complications among HTx patients. Our aim was to provide an update for the benefit/risk profile for surveillance and for cause EMBs.

## Methods

### Data Sharing

The data that support the findings of this study are openly available in Mendeley Data (doi:10.17632/vyrdvb8fv9.1).

### Study Design

This study was a retrospective, observational study of consecutive EMBs performed on HTx patients at the University of California, San Diego Health (UC San Diego Health) between August 2019 to August 2022. Eligible patients were HTx recipients who were 18 years of age or older who survived to their first EMB. All EMBs at UC San Diego Health are fluoroscopy-guided and at least 3 separate passes for EMB samples are attempted.^16^ For this study, the authors (VC, NR, BG) extracted patient data and clinical outcomes from the electronic medical record. Estimated glomerular filtration rate was calculated using the chronic kidney disease epidemiology collaboration equation, which does not include a race factor.^17^ Approval for this study was provided by the UC San Diego Health Office of IRB Administration (#805675). This study adheres to the principles of the Declaration of Helsinki formulated by the World Medical Association and the US Federal Policy for the Protection of Human Subjects.

### EMB Complications

Potential EMB complications included: new pericardial effusions that required intervention, new pericardial effusions moderate or greater in size that was increased by more than 1 grade, tricuspid valve injury, inadvertent arterial access, failed venous access attempts, atrial or ventricular arrhythmias, atrioventricular block, pneumothorax, hemothorax, access site infection or hematoma, arteriovenous fistula, pseudoaneurysm, vasovagal reaction, and coronary artery fistula. To meet criteria for a procedure-related pericardial effusion, there had to be prior cardiac imaging by echocardiography or computed tomography for comparison to demonstrate that the pericardial effusion was a new finding after the procedure. To meet criteria for a procedure-related tricuspid valve injury, there had to be a prior echocardiogram for comparison with a new diagnosis of moderate or greater tricuspid valve regurgitation that was increased by more than 1 grade and was found to be persistent in subsequent echocardiograms.^5^ All EMB complications were adjudicated by two experienced HTx cardiologists (NW and PJK). Where there was disagreement, a third cardiologist (YT) made the final determination.

### For Cause vs Surveillance

At UC San Diego Health, surveillance EMB are performed biweekly for the first 3 months and monthly afterwards. After 1 year post-HTx, surveillance EMBs are no longer performed except in rare cases where a patient is deemed high risk for recurrent AR. For cause refers to an EMB performed for clinical suspicion of rejection which includes: signs or symptoms of congestive heart failure, echocardiographic evidence of graft dysfunction (left ventricular ejection fraction < 50%), new arrhythmias, repeat EMB requested to confirm the resolution of a recent episode of AR, and development of a de novo donor specific antibody (DSA). EMBs performed with concurrent but not de novo DSA were considered surveillance unless there was documentation indicating clinical suspicion for rejection.

### Biopsy-defined Rejection

We followed the ISHLT classification scheme for clinically significant acute cellular rejection (ACR) and antibody mediated rejection (AMR). AR refers to either clinically significant ACR, AMR, or both (mixed ACR and AMR).^3^ At UC San Diego Health, a weekly pathologic review of all EMB samples is performed as previously described.^18^ Treatment for AR refers to a significant change in a subject’s immunomodulatory regimen including: initiation or increase in corticosteroids to a prednisone equivalent of 40 mg/day or higher, intravenous immune globulin, plasmapheresis, rituximab, thymoglobulin, and/or bortezomib use.

### Clinical Outcomes

All HTx patients were followed for all-cause death. Cause of death was adjudicated by NW, PJK and YT. Additional days of hospitalization after an EMB complication refers to the number of days beyond the initial projected hospitalization discharge date.

### Statistical Analyses

Categorical variables were expressed as frequency and percentage and compared with the use of either the Pearson’s chi-square or Fisher’s exact test. Continuous variables were expressed as mean ± standard deviation (SD) for normally distributed variables or median and interquartile range (IQR) for non-normally distributed variables and compared with the use of the Student’s t-test or Wilcoxon rank-sum test as appropriate.

We calculated the prevalence of EMB complications, AR, and treated AR and compared differences in proportions between the surveillance and for cause groups. The agreement rate of the initial adjudication for EMB complications was analyzed by Cohen’s kappa statistics. For EMB complications, treated AR, and to identify candidate predictors for prediction models, we performed mixed effects logistic regression with forward model selection to take into account within-subject correlation and determine significant predictors at a subject level using a p-value less than 0.15 threshold. The benefit/risk ratio was calculated simply by N of treated AR versus N of EMB complications. No weighting of treated AR was performed due to the uncertain benefit of treating asymptomatic 2R ACR and AMR rejections.^19, 20^ Poisson models were used to evaluate the benefit/risk ratios in the for cause compared to surveillance groups via an interaction term to account for within-subject correlation.

Analysis was conducted in R (R Core Team, 2022). We used the Bonferroni-Holm procedure whenever multiple comparisons were performed while implementing a particular statistical hypothesis test. The corrected p values are designated as p_c_. For single hypothesis testing we report the unadjusted p value. P or p_c_ < 0.05 are considered significant.

## Results

### Characteristics of the study population

A total of 2,769 consecutive EMBs from 326 unique HTx patients were included in this study. All cases were included for the primary outcome of EMB complications (**Figure 1**). For cause EMBs accounted for 499 (18.0%) samples while surveillance EMBs accounted for 2,270 (82.0%) samples.

**Figure 1.**
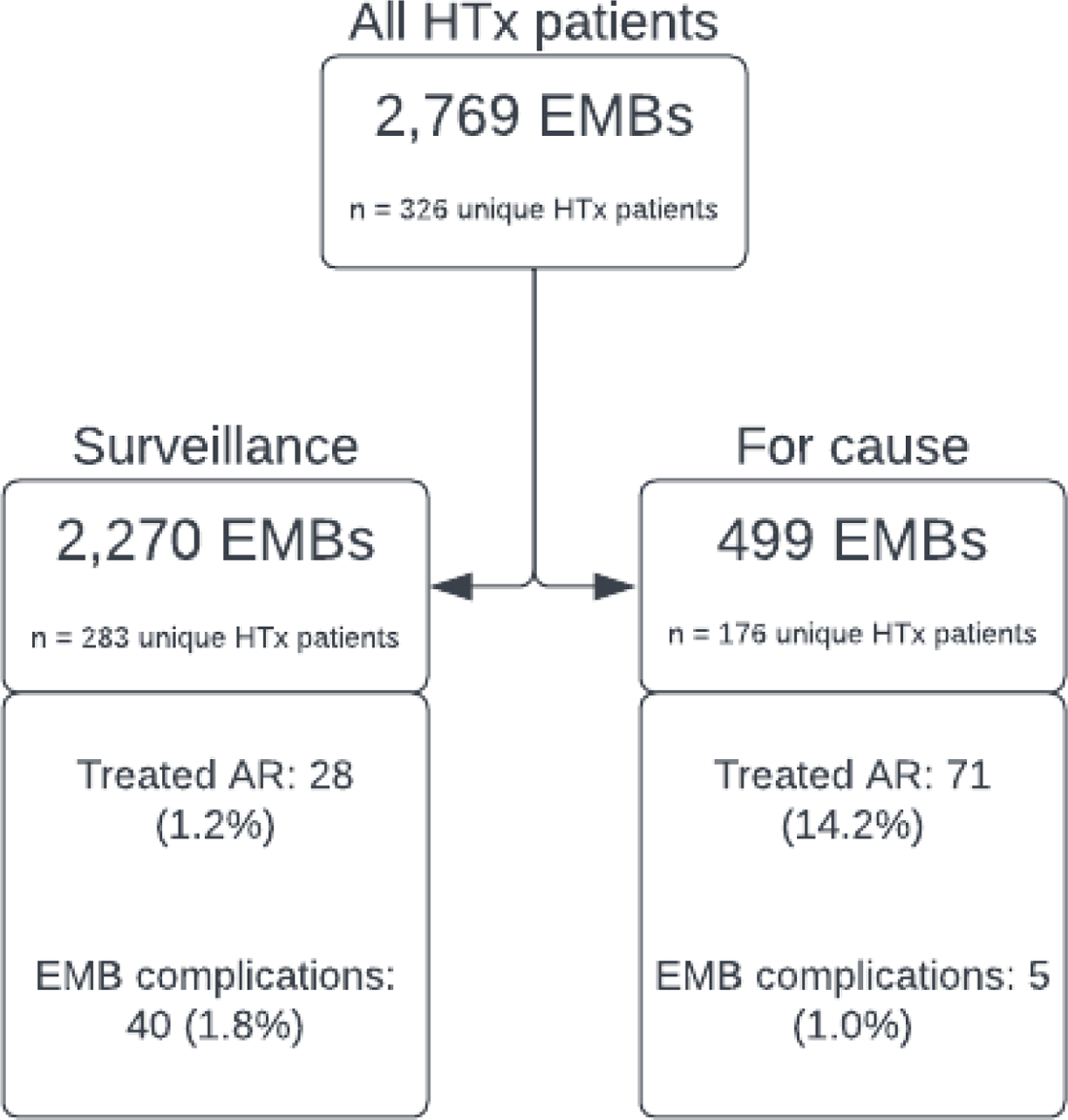
Flow diagram. AR, acute rejection; EMB, endomyocardial biopsy

Baseline characteristics of the study population are depicted in **Table 1**. Patients were typically male (78.8%) and non-hispanic white (39.0%) with a mean age of 55.5 ± 13.8 years. There were 944.8 person-years in this study from HTx to end of follow-up.

**Table 1.** Characteristics of Subjects and Endomyocardial Biopsies. DCD, donation after cardiac death; DPP-NMP, direct procurement perfusion-normothermic machine perfusion; ICM, ischemic cardiomyopathy; NICM, nonischemic cardiomyopathy; NRP-CSS, normothermic regional perfusion-cold static storage; PHM, predicted heart mass; PRA, panel reactive antibodies; VAD, ventricular assist device

|  | N |  |
| --- | --- | --- |
| **Donor characteristics** |  |  |
| Age, y, mean (SD) | 321 | 33.3 (10.7) |
| Male, N (%) | 326 | 267 (81.9) |
| **Recipient characteristics** |  |  |
| Age, y, mean (SD) | 326 | 55.5 (13.8) |
| Male, N (%) | 326 | 257 (78.8) |
| Race |  |  |
| Asian, N (%) | 326 | 24 (7.4) |
| Black, N (%) | 326 | 44 (13.5) |
| Native American, N (%) | 326 | 2 (0.6) |
| Other Race, N (%) | 326 | 27 (8.3) |
| Pacific Islander, N (%) | 326 | 4 (1.2) |
| White, N (%) | 326 | 225 (69.0) |
| Ethnicity |  |  |
| Hispanic or Latino, N (%) | 326 | 98 (30.1) |
| Not Hispanic or Latino, N (%) | 326 | 228 (69.9) |
| **Transplant characteristics** |  |  |
| Multi-organ transplant, N (%) | 326 | 53 (16.3) |
| Total donor ischemic time, min, mean (SD) | 319 | 211 (70) |
| Sex mismatch, N (%) | 326 | 54 (16.6) |
| PHM difference, % recipient PHM, mean (SD) | 321 | 5.9 (21.7) |
| Sensitized patients (PRA ≥ 10%) | 319 | 58 (18.2) |
| VAD use, N (%) | 326 | 108 (33.1) |
| Indication for Transplant |  |  |
| NICM, N (%) | 326 | 188 (57.7) |
| ICM, N (%) | 326 | 113 (34.7) |
| Congenital, N (%) | 326 | 18 (5.5) |
| Retransplant, N (%) | 326 | 7 (2.1) |
| Induction therapy |  |  |
| Thymoglobulin, N (%) | 326 | 109 (33.4) |
| Basiliximab, N (%) | 326 | 26 (8.0) |
| Eculizumab, N (%) | 326 | 2 (0.6) |
| DCD, N (%) | 326 | 65 (19.9) |
| NRP-CSS | 326 | 49 (15.0) |
| DPP-NMP | 326 | 16 (4.9) |
| **Endomyocardial biopsy characteristics** |  |  |
| Time post-transplant, d, median (IQR) | 2769 | 100 (48-217) |
| De novo DSA, N (%) | 2757 | 74 (2.7) |
| Concurrent DSA, N (%) | 2757 | 233 (8.5) |
| Concurrent cardiac allograft dysfunction, N (%) | 2769 | 135 (4.9) |

EMB procedural characteristics are summarized in **Table S1**. All EMBs were performed using fluoroscopy-guidance and the majority were performed using the right internal jugular vein as access (84.5%). Eight different HTx cardiologists performed EMBs for this study. The median number of samples per EMB was 4 (IQR, 4-5). The median number of EMB per patient was 9 (IQR, 6-12). Most EMBs were performed in an outpatient setting (78.8%).

### EMB complications

In the study population, 45 (1.6%) total complications occurred in 41 unique HTx patients. Initial adjudication of EMB complications was in agreement 90.6% of the time with a Cohen’s kappa of 0.81 (0.64, 0.97; p < 0.001). There were 33 (73.3%) clinically significant pericardial effusions and 26 of the 33 pericardial effusions required a percutaneous or surgical intervention, for an effusion of moderate or larger size not previously observed by echocardiography. Other complications were less frequent and are shown in **Table 2**. There was a mean of 4.0 (95% CI, 2.89-5.11; p < 0.001) additional days of hospitalization due to an EMB complication. Clinically significant pericardial effusions occurred separately twice in two HTx patients and no HTx patient had more than two EMB complications. There were 11 (0.4%) non-diagnostic EMB samples in our study. Repeat EMB was performed in 7 of the 11 non-diagnostic cases. No repeat EMBs were associated with complications and 4 of the 7 repeat EMBs were performed in the first month after HTx.

**Table 2.** Endomyocardial biopsy complications.

|  | Endomyocardial cases | Unique heart transplant patients |
| --- | --- | --- |
| Clinically significant pericardial effusion – no. of patients/total no. (%) | 33/45 (73.3) | 31/41 (75.6) |
| Pericardiocentesis with pericardial drain – no. of patients/total no. | 25/33 | 24/31 |
| Surgical pericardial window – no. of patients/total no. | 1/33 | 1/31 |
| Tricuspid valve injury – no. of patients/total no. (%) | 3/45 (6.7) | 3/41 (7.3) |
| Inadvertent arterial access – no. of patients/total no. (%) | 3/45 (6.7) | 3/41 (7.3) |
| Failed venous access attempt – no. of patients/total no. (%) | 4/45 (8.9) | 4/41 (9.8) |
| Right atrial lead dislodgement – no. of patients/total no. (%) | 1/45 (2.2) | 1/41 (2.4) |
| Extraction of embedded biptome – no. of patients/total no. (%) | 1/45 (2.2) | 1/41 (2.4) |

We evaluated 47 predictors for EMB complications (**Table S2**) using single predictor logistic regression. Using multi-predictor logistic regression, only time since HTx was found to be a significant predictor for EMB complications with the highest risk period to be within 1 month after HTx (OR 12.74; 95% CI, 6.67-24.40; p_c_ < 0.001; **Figures 2A and 2B**). There was a nonsignificant trend for increased EMB complications with surveillance indication (p = 0.230). Other factors including bioptome size, different operators, trainee involvement, and elevated intracardiac filling pressures were not found to be significantly associated with EMB complications after adjusting for multiple covariates.

**Figure 2.**
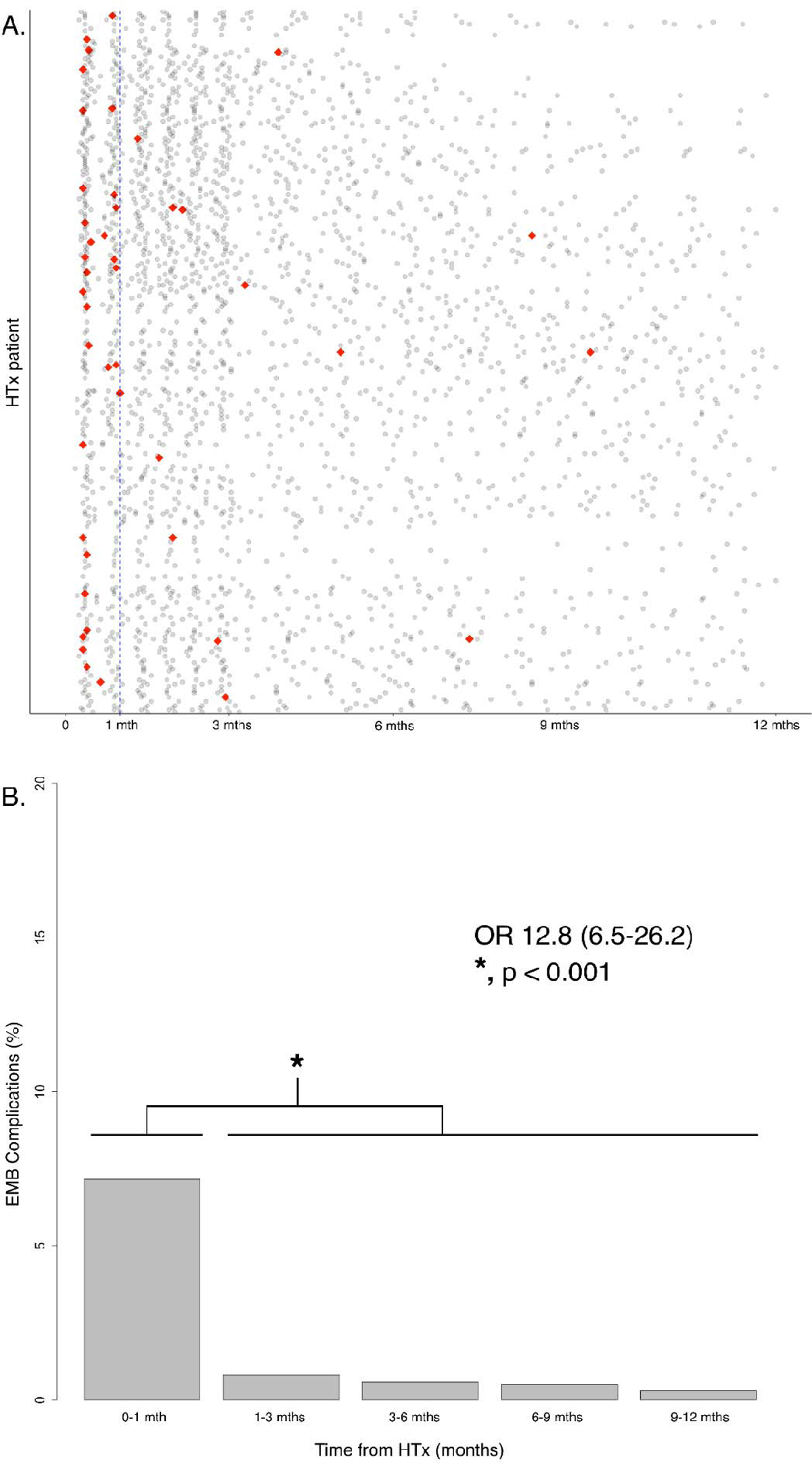
Endomyocardial biopsy (EMB) complications over time since heart transplantation (HTx). **A.** Scatterplot of EMBs for all 326 HTx patients. Each gray dot represents an EMB sample negative for EMB complications and each red diamond represents an EMB sample associated with a complication. EMB complications show a pattern of occurring within the first month after HTx. **B**. Barplot showing percentage of EMB complications within each time interval. There is a significant difference in percentage of EMB complications occurring in the first month compared to the rest of the first year after HTx (p < 0.001).

The rate of significant pericardial effusions, defined as pericardial effusions moderate or greater in size, was low at 1.7%. We found no significant association for donor-recipient predicted heart mass (PHM) mismatch (i.e., small donor heart transplanted in a large HTx recipient) and incidental pericardial effusion (p = 0.120). The majority of pericardial effusions were adjudicated as EMB complications (67.3%; 95% CI, 52.3%-79.6%). While ACR was not associated with pericardial effusions, AMR demonstrated a significant correlation with incidental pericardial effusions (OR 3.63; 95% CI, 1.39-9.49; p = 0.009). However, AMR did not significantly correlate with pericardial effusions that were adjudicated as EMB complications (p = 0.725).

Sensitivity analysis with EMB-related pericardial effusion as the outcome was also performed. Only EMBs performed within 1 month after HTx (OR 43.25; 95% CI, 14.91-125.50; p_c_ < 0.001) were found to be significantly associated with EMB-related pericardial effusion.

### Treated AR by EMB

AR was diagnosed in 133 (4.8%) EMB samples from 67 (20.6%) unique HTx patients (**Table S3**). However, only 99 (3.6%) AR samples from 61 (18.7%) unique HTx patients were treated. There was one EMB sample negative for ACR and AMR that was treated in the setting of focal myocyte necrosis and inflammation and concurrent cardiac allograft dysfunction. All untreated samples showed AMR without ACR (i.e., ACR 0R or 1R grades). Of the 35 untreated AMR samples, 28 were pAMR1 and 7 were pAMR2, including 2 patients that recently received immunomodulatory therapy and 1 patient that refused treatment.

We found treated AR diagnosed more frequently in for cause samples (14.2%) compared to surveillance EMB samples (1.2%; p < 0.001). The for cause indication demonstrated a significantly increased OR of 9.17 (95% CI, 4.56-18.46; p_c_ < 0.001; **Table S4**) for the diagnosis of treated AR. We found time from HTx was not significantly associated with treated AR after adjusting for multiple covariates (p_c_ = 0.909; **Figure 3**). We did not observe a significantly increased time interval between EMBs for treated AR samples compared to samples without treated AR (3.7 ± 2.4 versus 3.4 ± 2.1 weeks; p = 0.300).

**Figure 3.**
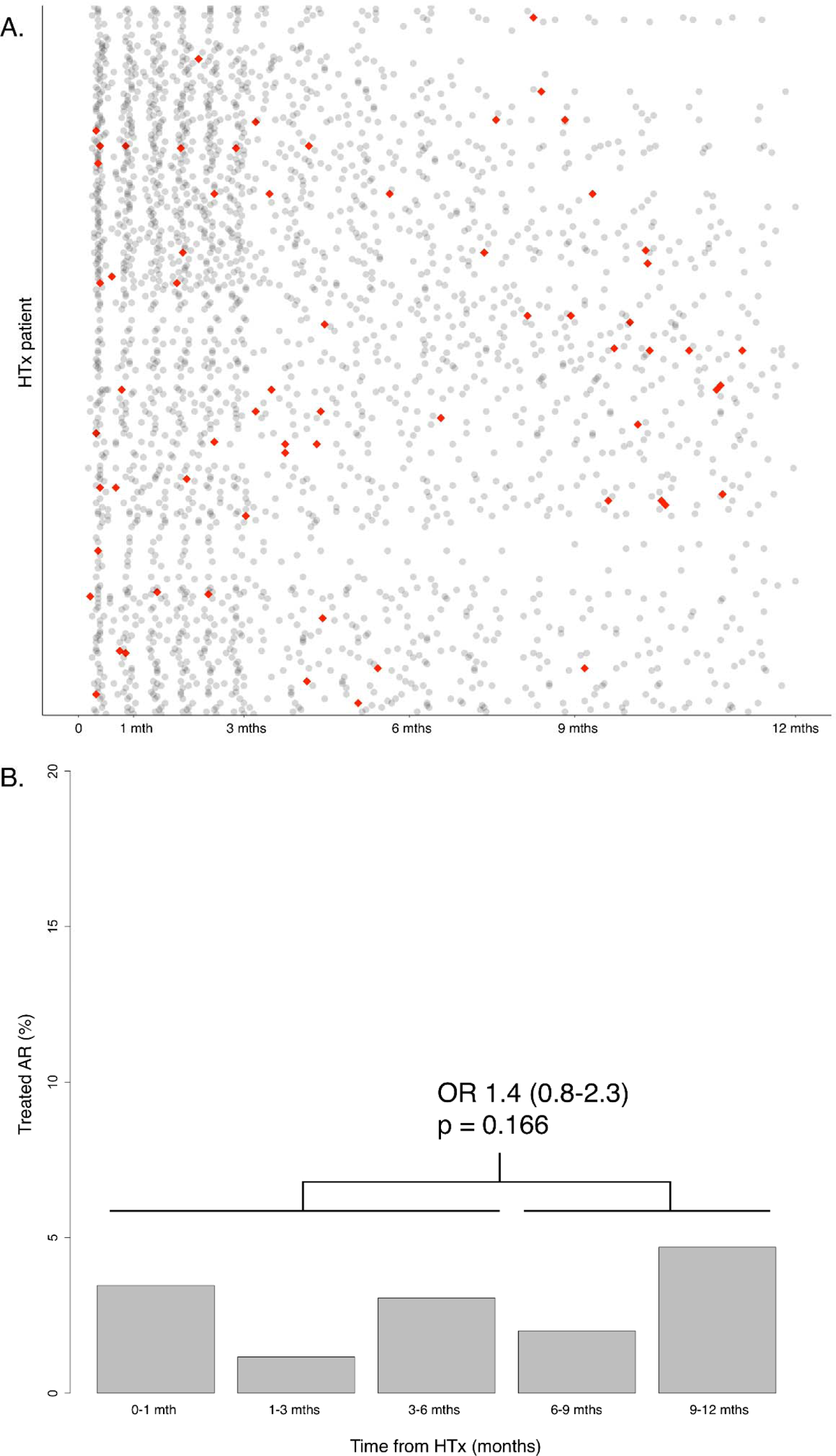
Treated acute rejection (AR) over time since heart transplantation (HTx). **A**. Scatterplot of endomyocardial biopsies (EMB) for all 326 HTx patients. Each gray dot represents an EMB sample negative for treated AR and each red diamond represents an EMB sample positive for treated AR. **B**. Barplot showing percentage of treated AR within each time interval. There is no significant difference in the percentage of treated AR in 0-6 months compared to 6-12 months after HTx (p = 0.17).

**Figure 4.**
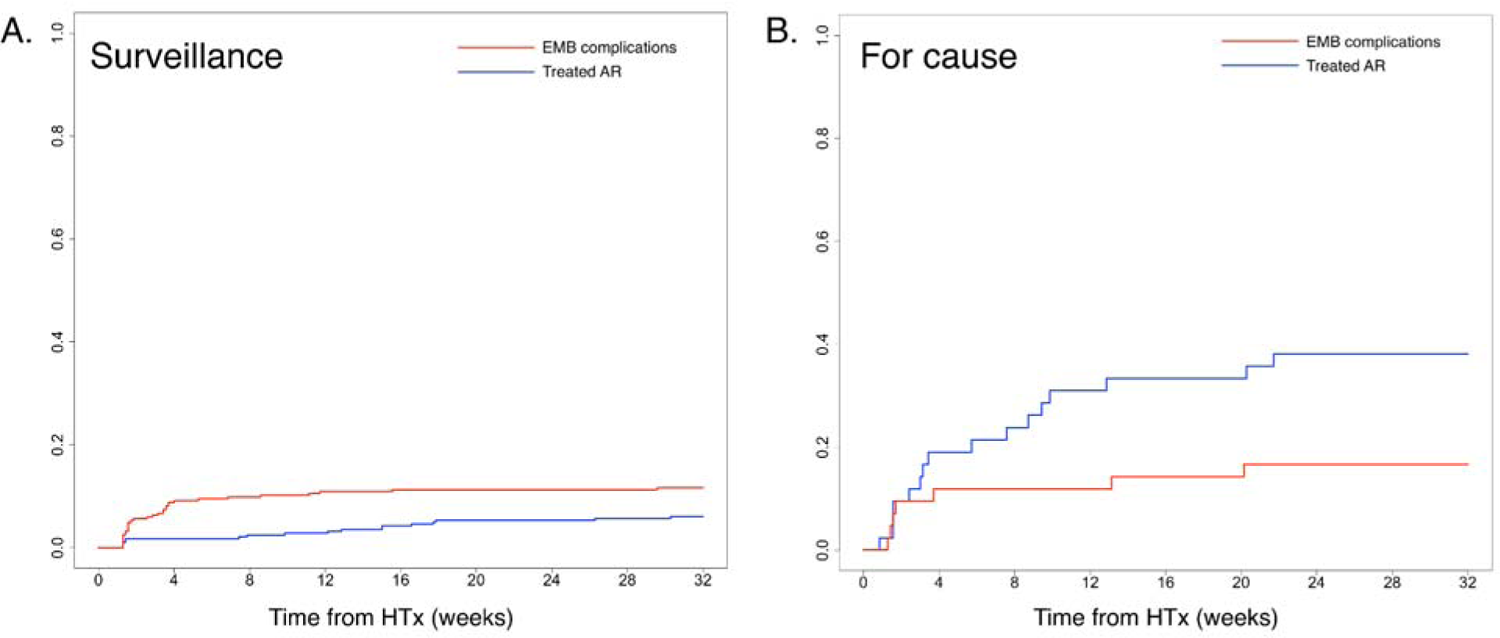
Endomyocardial biopsy complication (EMB) and treated acute rejection (AR) over time since heart transplantation (HTx). **A**. Surveillance EMB cumulative incidence curves for benefit and risk. EMB complications incidence acutely increases within the first month after HTx. Incidence of treated AR does not increase above the rate of EMB complications for surveillance EMBs. **B**. For cause EMB cumulative incidence curves for benefit and risk. The incidence of treated AR increases above the rate of EMB complications in for cause EMBs within the first month after HTx.

Of the EMB samples within 1 month after HTx, 382 (88.2%) were surveillance and 51 (11.8%) were for cause (**Table S5**). For this study, if an EMB was previously scheduled as a surveillance EMB but the clinical team noted concerns for possible AR prior to the procedure, the EMB was recategorized as for cause EMB. Of the EMB samples after 1 month HTx, 1,888 (80.8%) were surveillance and 448 (19.2%) were for cause. The number of EMB samples performed for cause was significantly increased after 1 month HTx compared to surveillance (OR 1.78; 95% CI, 1.30-2.47; p = 0.002). Among surveillance EMB within 1 month after HTx, we found 5 out of 11 AR samples prompted treatment. The 6 surveillance AR cases that were not treated were all pAMR1 without concurrent DSA. Among the for cause EMB within 1 month after HTx, we found 10 out of 11 AR samples were treated and the 1 untreated AR sample was a pAMR1 (I+) without concurrent DSA that was subsequently followed by repeated EMB until resolution of the AMR.

### Benefit of detecting treated AR compared to risk of EMB complications in for cause versus surveillance EMBs

The overall benefit/risk ratio (i.e., treated AR/EMB complication) was 2.2. In the for cause EMB group, we found the benefit/risk ratio increased to 14.2. In contrast, we found the benefit/risk ratio decreased to 0.7 in the surveillance EMB group. As a result, the ratio of benefit/risk ratios comparing surveillance to for cause EMB groups was significantly decreased at 0.05 (p < 0.001).

### Benefit of detecting treated AR compared to risk of EMB complications in EMBs performed before and after 1 month from HTx

We found the benefit/risk ratio was significantly improved in surveillance EMB when comparing EMBs performed after 1 month from HTx versus within 1 month after HTx (OR 11.59; 95% CI, 3.28-49.31; p < 0.001). However, we did not observe the benefit/risk ratio in for cause EMBs to be significantly different when comparing EMBs performed after 1 month from HTx versus within 1 month after HTx (OR 3.96; 95% CI, 0.30-39.39; p = 0.200).

### Clinical Outcomes

There were 24 deaths (7.4%; **Table S6**) and 1 retransplant. The majority of the deaths (45.8%) were due to infection. Of the 11 deaths due to infection, 7 (63.6%) were on either triple or quadruple immunosuppression and 10 (90.9%) were still taking prednisone. We did not observe any treated AR episodes in the preceding EMB prior to the diagnosis of the fatal infection. AR accounted for 3 (12.5%) deaths and all were due to AMR. However, the AMR deaths were outside of the surveillance biopsy window (129.0 ± 33.1 weeks) and all 3 patients were noted to have a history of non-adherence. All AMR episodes were determined by for cause EMBs.

## Discussion

In this retrospective single-center study, several key findings were observed. First, the rate of treated AR compared to EMB complications, calculated as the benefit/risk ratio, was significantly lower in surveillance compared to for cause EMBs. Second, we found the highest risk period for EMB complications to be within 1 month after HTx. Third, in the contemporary era, the benefit of detecting treated AR has decreased to the extent that we found the risk for EMB complications outweighed the benefit in surveillance EMBs. The benefit/risk ratio for surveillance EMBs improved after 1 month HTx, not because of any increase in benefit but due to the significant decrease in EMB complications. Fourth, treatment of AMR is inconsistent in the contemporary era and almost half of AMR EMBs do not lead to a change in treatment.

We found the EMB complication rate to be low (1.6%) with rates similar to previous studies.^5, 6^ While not a direct cause of death, EMB complications did contribute to increased morbidity, additional interventions, and a significant increase in time hospitalized by 4 days per EMB complication. Historically, tricuspid valve injury and vascular complications made up a significant portion of EMB complications.^6, 21^ However, pericardial effusions contributed to a greater proportion of EMB complications in more recent literature, consistent with our study findings.^5, 6, 8, 22^ We hypothesize that vascular complications and tricuspid valve injury have decreased due to improved techniques utilizing ultrasound for vascular access and increased attention to avoiding tricuspid valve injury, respectively.^23^ In contrast, the incidence of pericardial effusions as an EMB complication is likely unchanged due to the fact that the majority of studies, including this study, continue to report the practice of fluoroscopy-guided EMBs.^5, 6, 8^ However, despite the theoretical benefit of echo-guided EMBs, no studies have have demonstrated a significant decrease in EMB complications with the use of echo-guided compared to fluoroscopy-guided EMBs.^24, 25^ At our institution, fluoroscopy-guided EMBs are solely performed due to provider preference and lack of sonographers with the expertise to guide HTx cardiologists in performing echo-guided EMBs. These reasons also likely explain why fluoroscopy-guidance will continue to remain the predominant method for EMBs for most institutions.^5, 9^ Compared to other studies, our patients more frequently prompted intervention for the pericardial effusion, which likely reflect differences in practice patterns. Furthermore, we did not find that donor heart size contributed to the development of incidental pericardial effusion, which itself was a rare event.

Our study showed a novel finding that earlier time from HTx was associated with a higher rate of EMB complications, with the rate significantly increased within the first month after HTx compared to after 1 month from HTx. This finding was driven by a significantly increased rate of EMB-related pericardial effusions. We hypothesize that myocyte necrosis from ischemia-reperfusion injury and its persistence related to immunosuppression predisposes patients to EMB-related pericardial effusions within 1 month after HTx.^26, 27^ This hypothesis is also supported by greater levels dd-cfDNA early in the post-HTx period, indicating a vulnerable period due to allograft injury in the early post-HTx period.^2^ While prior studies have described the rate of EMB complications, this is the first study to identify a potential risk factor associated with EMB complications. This finding has significant clinical relevance and future studies should further evaluate this high risk time period to identify potential strategies to reduce the risk of EMB complications.

Our study findings also corroborated a reduced incidence of ACR in the contemporary era compared to prior eras, attributed to modern immunosuppression regimens and improved post-HTx care.^11, 12, 28^ In contrast to earlier eras,^6, 28^ our findings showed that time from HTx was not independently associated with treated ACR.^22^ This is an important observation that suggests that time from HTx is no longer relevant as a risk factor for AR. Thus, similar to the current approach in pediatric HTx patients, our study findings suggest it may be reasonable to be guided by “clinical vigilance” for AR surveillance, regardless of the time interval from HTx.^15^ In addition to other literature,^8, 28^ our study findings suggest sufficient equipoise for future randomized controlled trials that include clinical vigilance as a study arm to determine whether surveillance EMBs are better than clinical assessment alone, a fact that has not yet been established in the contemporary era.^29^

In contrast to our study findings with respect to ACR, we observed an increased incidence of AMR, likely due to increasing awareness of AMR.^30, 31^ Although a large proportion of AMR (42.7%) were not treated, the absolute number of treated AMR was greater than ACR in our study. However, despite the increase in treated AMR, the total number and rate of treated AR remained low. The inconsistency in treatment of AMR reflects the current uncertainty of benefit with treatment.^32^ Until there is consensus in the HTx field for treating AMR, the benefit for asymptomatic AMR patients will likely remain low.

Our study demonstrates that for cause EMB still detects treated AR at a high rate and remains an important tool in the clinical armamentarium for HTx patients with signs or symptoms concerning for AR. In our study, we also demonstrated a trend towards reduced EMB complications in the for cause group as the utilization of for cause compared to surveillance EMBs was significantly decreased within 1 month after HTx. Thus, another possible benefit of prioritizing for cause EMBs would be reducing EMBs performed within 1 month after HTx, a high risk time period for EMB complications.

With the improvement of HTx care, AR rates have significantly decreased to the point where noninvasive biomarker tests have demonstrated both safety and non-inferiority to surveillance EMBs.^33^ IMAGE was the first and largest clinical trial that demonstrated the use of GEP could safely replace surveillance EMB as early as 6 months after HTx in low-risk patients. The eIMAGE trial expanded on this result to show that GEP could be safely used as early as 55 days after HTx, again in low-risk patients.^34^ However, a recurring critique for both studies was the lack of an appropriate “control” arm of clinical vigilance, given the low rate of pre-specified outcome events for both studies.^35^ Our study suggests that, in a real world setting that includes both low and high risk HTx patients in the contemporary era, surveillance EMB may no longer provide any clinical benefit without incurring a greater risk of cardiac injury in the early post HTx period. Currently, the leading candidate for non-invasive AR surveillance, dd-cfDNA, has limited accuracy in this high risk time period.^2, 18^ Further efforts to improve dd-cfDNA testing^3^ could be a potential solution to reduce the risk of cardiac injury while increasing the potential benefit by identifying HTx patients more likely to have AR. Benefit/risk of EMBs could be also improved by coming to a consensus for when to treat AMR and thus limiting EMBs to scenarios where the results can potentially change treatment. In addition, knowledge of this high risk time period for EMB complications may itself be useful in reducing EMB complications. Clinicians can use this important information to be more cautious during these higher risk EMBs to possibly reduce the rate of complications, similar to the reductions seen in vascular complications and tricuspid valve injuries. HTx programs may also re-evaluate the necessity of surveillance EMB during this time period. Finally, our study findings contribute to prior literature to suggest sufficient clinical equipoise for future randomized controlled trials to compare surveillance EMBs with “standardized clinical and functional allograft vigilance.”^8, 19, 28, 35, 36^ We believe these trials are necessary in the contemporary era due to decreasing benefit/risk ratio and would also complement the growing research in noninvasive biomarkers, including dd-cfDNA testing.

### Limitations

This study should be interpreted within the context of several important limitations. First, this was a retrospective study from a single center and may not necessarily represent the experience of other centers with different patient demographics, procedural characteristics, and variations in post-HTx management. Second, UC San Diego Health does not consistently perform echocardiograms after every EMB, as reported in some other studies.^5, 6^ However, the incidence of pericardial effusions in this study is similar to recent studies.^5, 22^ Third, all EMBs in this study were performed with fluoroscopic guidance. Increased use of echo-guidance could potentially further reduce EMB complications. Finally, the benefit/risk ratio gives equal weight to complications, including some that may be considered minor, and treatment of AR. Because the benefit of treating specific ARs, including asymptomatic 2R ACR and AMR episodes, is uncertain,^8, 19, 20^ we simply calculated the benefit/risk ratio using the N of AR vs N of EMB complications for consistency and objectivity.

## Conclusion

Detection of treated AR by surveillance EMBs in adult HTx patients has declined in the contemporary era resulting in a significantly lower benefit/risk ratio in surveillance compared to for cause EMBs. Future randomized controlled trials that compare surveillance EMBs to clinical assessment alone are necessary to evaluate whether our current practice of surveillance EMBs should be continued in the contemporary era.

## Acronyms

ACR: Acute cellular rejection

AMR: Antibody mediated rejection

AR: Acute rejection

dd-cfDNA: donor-derived cell-free

DNA DSA: donor specific antibody

EMB: endomyocardial biopsy

GEP: gene-expression profiling

HTx: heart transplant

LVEF: Left ventricular ejection fraction

IQR: interquartile range

pAMR: pathological antibody mediated rejection

PHM: predicted heart mass

SD: standard deviation

UC San Diego Health: University of California, San Diego Health

## Data Availability

The data that support the findings of this study are openly available in Mendeley Data at 10.17632/vyrdvb8fv9.1.

https://data.mendeley.com/datasets/cpyj7v99rj/2

## Acknowledgements

The authors acknowledge Melissa McLenon, DNP, ACNP, from UC San Diego Health for her contribution to data collection and Priyesha Bijlani, MD, from UC San Diego Health for her contribution to the review and editing of the final manuscript.

## Author contributions

VC: conceptualization, writing, review and editing, data curation, project administration; FV: writing, review and editing, statistical analyses; NW: writing, review and editing, data curation, adjudication of endomyocardial biopsy complications and clinical outcomes; NR: writing, review and editing, data curation; YT: writing, review and editing, data curation, adjudication of endomyocardial biopsy complications and clinical outcomes; Bryn Gerding: writing, review and editing, data curation; Barry Greenberg: writing, review and editing; MAU: writing, review and editing; EAdler: writing, review and editing; PJK: conceptualization, writing, review and editing, data curation, adjudication of endomyocardial biopsy complications and clinical outcomes, project administration.

## Acknowledgements/Funding

This study was supported by the Altman Clinical & Translational Research Institute (ACTRI) at UC San Diego Health (PJK). The ACTRI is funded from awards issued by the National Center for Advancing Translational Sciences, NIH KL2TR001444 and NIH UL1 TR001442. Dr. Nicholas Wettersten and this work was supported (or supported in part) by Career Development Award Number IK2 CX002105 from the United States (U.S.) Department of Veterans Affairs Clinical Sciences R&D (CSRD) Service. The contents do not represent the view of the U.S. Department of Veterans Affairs or the United States Government.

