## Supplemental Tables for "Benefit versus Risk of Endomyocardial Biopsy for Heart Transplant Patients in the Contemporary Era"

**SUPPLEMENTARY RESULTS**

**Table S1. Additional characteristics of subjects and endomyocardial biopsies.** ACR, acute cellular rejection; AMR, antibody mediated rejection; BUN, blood urea nitrogen; DOAC, direct oral anticoagulant; DPP-NMP, direct procurement perfusion-normothermic machine perfusion; ECMO, extracorporeal membrane oxygenation; HTX, heart transplant; IABP, intra-aortic balloon pump; LPM, liters per minute; MCS, mechanical circulatory support; NRP-CSS, normothermic regional perfusion-cold static storage; UNOS, United Network of Organ Sharing

| Variables | N | Value |
| --- | --- | --- |
| **Recipient characteristics** | | |
| Recipient body mass index, kg/m^2^, mean (SD) | 326 | 26.3 (4.5) |
| **Donor characteristics** |  |  |
| Donor body mass index, kg/m^2^, mean (SD) | 309 | 27.2 (5.9) |
| **Transplant characteristics** | | |
| Primary graft dysfunction | 326 | 61 (18.7) |
| UNOS status | | |
| Status 1 (post 10/2018), N (%) | 316 | 10 (3.2) |
| Status 2 (post 10/2018), N (%) | 316 | 99 (31.3) |
| Status 3 (post 10/2018), N (%) | 316 | 45 (14.2) |
| Status 4 (post 10/2018), N (%) | 316 | 81 (25.6) |
| Status 5 (post 10/2018), N (%) | 316 | 13 (4.1) |
| Status 6 (post 10/2018), N (%) | 316 | 33 (10.4) |
| Status 1A (pre 10/2018), N (%) | 316 | 23 (7.3) |
| Status 1B (pre 10/2018), N (%) | 316 | 10 (3.2) |
| Status 2 (pre 10/2018), N (%) | 316 | 2 (0.6) |
| Temporary MCS pre-transplant | | |
| ECMO, N (%) | 321 | 8 (2.5) |
| IABP, N (%) | 321 | 91 (28.4) |
| Temporary MCS post-transplant | | |
| ECMO, N (%) | 321 | 16 (5.0) |
| IABP, N (%) | 321 | 51 (15.9) |
| NRP-CSS | | |
| Total ischemic time, min, median (IQR) | 49 | 236.0 (204.0-273.0) |
| Functional warm ischemic time, min, median (IQR) | 49 | 24.0 (20.0-37.0) |
| Cold ischemic time, min, median (IQR) | 49 | 178.0 (147.0-230.0) |
| Warm ischemic time, min, median (IQR) | 49 | 48.0 (43.0-51.0) |
| DPP-NMP | | |
| Total ischemic time, min, median (IQR) | 16 | 98.0 (87.5-109.5) |
| Functional warm ischemic time, min, median (IQR) | 16 | 24.5 (18.0-29.2) |
| Cold ischemic time, min, median (IQR) | 16 | 44.0 (41.0-51.5) |
| Warm ischemic time, min, median (IQR) | 16 | 48.5 (44.8-53.2) |
| **Endomyocardial biopsy characteristics** | | |
| For cause indication | 2769 | 499 (18.0) |
| Cardiac allograft vasculopathy, N (%) | 224 | 14 (6.3) |
| Dialysis, N (%) | 2769 | 36 (1.3) |
| History of ACR, N (%) | 2769 | 334 (1.2) |
| History of AMR, N (%) | 2769 | 360 (13.0) |
| Pericardial effusion, N (%) | 2769 | 49 (1.8) |
| Trainee involved, N (%) | 2769 | 855 (30.9) |
| Outpatient status, N (%) | 2769 | 2182 (78.8) |
| Fluoroscopy time, min, mean (SD) | 2624 | 3.3 (3.3) |
| Combined with left heart catheterization | 2769 | 5 (0.2) |
| Number of biopsy samples, median (IQR) | 2762 | 4.0 (4.0-5.0) |
| Venous access site | | |
| Femoral, N (%) | 2762 | 103 (3.7) |
| Right internal jugular, N (%) | 2762 | 2333 (84.5) |
| Left internal jugular, N (%) | 2762 | 288 (10.4) |
| Right brachial, N (%) | 2762 | 38 (1.4) |
| Bioptome size | | |
| 7 French, N (%) | 2759 | 1506 (54.6) |
| 6 French, N (%) | 2759 | 1017 (36.9) |
| 5.5 French, N (%) | 2759 | 236 (8.6) |
| Anticoagulant use, N (%) | 2769 | 345 (12.5) |
| DOAC, N (%) | 345 | 327 (94.8) |
| Warfarin, N (%) | 345 | 17 (4.9) |
| Heparin infusion, N (%) | 345 | 1 (0.3) |
| Right heart catheterization | | |
| Right atrial pressure, mmHg, mean (SD) | 1827 | 6.3 (4.1) |
| Pulmonary arterial pressure, mmHg, mean (SD) | 1825 | 20.4 (6.0) |
| Pulmonary capillary wedge pressure, mmHg, mean (SD) | 1805 | 11.7 (5.1) |
| Fick cardiac index, LPM/m^2^, mean (SD) | 1824 | 3.2 (0.8) |
| Laboratory values | | |
| BUN, mg/dL, mean (SD) | 2733 | 31.1 (17.9) |
| Creatinine, mg/dL, mean (SD) | 2732 | 1.4 (0.6) |
| Hemoglobin, g/dL, mean (SD) | 2769 | 11.3 (2.0) |
| Platelets, 1000/mm^3^, mean (SD) | 2769 | 248 (90.5) |

**Table S2. Single predictor logistic regression for endomyocardial biopsy complications**. ACR, acute cellular rejection; AMR, antibody mediated rejection; BMI, body mass index; BUN, blood urea nitrogen; CI, confidence interval; DPP-NMP, direct procurement perfusion-normothermic machine perfusion; DSA, donor specific antibody; ECMO, extracorporeal membrane oxygenation; eGFR, estimated glomerular filtration rate; EMB, endomyocardial biopsy; HTx, heart transplant; IABP, intra-aortic balloon; NRP-CSS, normothermic regional perfusion-cold static storage; OR, odds ratio; PHM, predicted heart mass; UNOS, United Network Organ Sharing.

| Variables | No. of EMBs | Total EMB complications | No. of patients with EMB complications | OR | 95% CI | p-value |
| --- | --- | --- | --- | --- | --- | --- |
| **Recipient characteristics** | | | | | | |
| Age (per 1-y increment) | 2769 | 45 | 41 | 1.01 | [0.99-1.03] | p = 0.390 |
| Male sex | 2769 | 45 | 41 | 1.63 | [0.72-3.73] | p = 0.243 |
| Race and ethnicity | 2769 | 45 | 41 | – | – | p = 0.168 |
| Recipient BMI (per kg/m^2^) | 2769 | 45 | 41 | 1.01 | [0.94-1.08] | p = 0.813 |
| Multi-organ transplant | 2769 | 45 | 41 | 1.25 | [0.58, 2.66] | p = 0.571 |
| HTx indication | 2769 | 45 | 41 | – | – | P = 0.250 |
| **Donor characteristics** | | | | | | |
| Age (per 1-y increment) | 2752 | 45 | 41 | 0.99 | [0.96-1.02] | p = 0.468 |
| Male sex | 2769 | 45 | 41 | 0.84 | [0.39-1.80] | p = 0.655 |
| Donor BMI (per kg/m^2^) | 2752 | 45 | 41 | 0.96 | [0.91-1.02] | p = 0.178 |
| **Transplant characteristics** | | | | | | |
| UNOS status | 2734 | 45 | 41 | – | – | p = 0.389 |
| Induction therapy | 2769 | 45 | 41 | – | – | p = 0.377 |
| Total donor ischemic time | 2735 | 45 | 41 | 1.00 | [1.00-1.00] | p = 0.967 |
| PHM difference | 2752 | 45 | 41 | 0.99 | [0.97-1.01] | p = 0.208 |
| Primary graft dysfunction | 2769 | 45 | 41 | 1.63 | [0.83-3.18] | p = 0.154 |
| Donation after cardiac death | 2769 | 45 | 41 | 0.75 | [0.34-1.64] | p = 0.471 |
| DPP-NMP | 2769 | 45 | 41 | 0.34 | [0.05-2.55] | p = 0.294 |
| NRP-CSS | 2769 | 45 | 41 | 0.90 | [0.39-2.08] | p = 0.812 |
| ECMO pre-HTx | 2748 | 45 | 41 | 0.70 | [0.09-5.33] | p = 0.727 |
| ECMO post-HTx | 2748 | 45 | 41 | 2.11 | [0.71-6.28] | p = 0.178 |
| IABP pre-HTx | 2748 | 45 | 41 | 1.88 | [1.02-3.44] | p = 0.042 |
| IABP post-HTx | 2748 | 45 | 41 | 1.15 | [0.54-2.45] | p = 0.720 |
| **Endomyocardial biopsy characteristics** | | | | | | |
| Within 1 month of HTx | 2769 | 45 | 41 | 13.05 | [6.82-24.97] | p < 0.001 |
| Time since HTx (per week) | 2769 | 45 | 41 | 0.92 | [0.89-0.96] | p < 0.001 |
| Surveillance indication | 2769 | 45 | 41 | 1.78 | [0.69-4.56] | p = 0.229 |
| Bioptome size | 2759 | 39 | 36 | – | – | p = 0.371 |
| Venous access site | 2762 | 42 | 38 | – | – | p = 0.177 |
| Fluoroscopy time (per min) | 2624 | 39 | 37 | 1.05 | [0.99-1.11] | p = 0.104 |
| Concurrent graft  dysfunction | 2769 | 45 | 41 | 0.91 | [0.21-3.87] | p = 0.899 |
| Concurrent DSA | 2758 | 45 | 41 | 0.49 | [0.12-2.08] | p = 0.336 |
| De novo DSA | 2757 | 45 | 41 | 1.72 | [0.40-7.31] | p = 0.460 |
| Number of EMB samples | 2762 | 39 | 36 | 1.01 | [0.74-1.39] | p = 0.935 |
| Anticoagulant use | 2769 | 45 | 41 | 1.08 | [0.45-2.60] | p = 0.865 |
| Hemodialysis | 2769 | 45 | 41 | 3.48 | [0.75-16.07] | p = 0.110 |
| Trainee involvement | 2769 | 45 | 41 | 1.83 | [1.00-3.33] | p = 0.048 |
| ACR | 2752 | 38 | 35 | – | – | p = 0.614 |
| AMR | 2754 | 38 | 35 | – | – | p = 0.484 |
| Inpatient status | 2769 | 45 | 41 | 4.04 | [2.22-7.34] | p < 0.001 |
| Combined with left heart  catheterization | 2769 | 45 | 41 | – | – | p = 1.0 |
| Right heart catheterization | | | | | | |
| Right atrial pressure (per 1 mmHg) | 1827 | 37 | 35 | 1.11 | [1.04-1.19] | p = 0.001 |
| Pulmonary artery pressure (per 1 mmHg) | 1825 | 37 | 35 | 1.05 | [1.00-1.10] | p = 0.066 |
| Pulmonary capillary wedge  pressure (per 1 mmHg) | 1805 | 35 | 34 | 1.09 | [1.03-1.16] | p = 0.002 |
| Fick cardiac index (per 1  LPM/m^2^) | 1824 | 37 | 35 | 1.10 | [0.74-1.64] | p = 0.639 |
| Laboratories | | | | | | |
| BUN (per 1 mg/dL) | 2733 | 43 | 39 | 1.02 | [1.01-1.03] | p < 0.001 |
| Creatinine (per 1 mg/dL) | 2732 | 43 | 39 | 1.19 | [0.80-1.78] | p = 0.393 |
| eGFR (per 1 mL/1.73 m^2^) | 2769 | 45 | 41 | 0.98 | [0.96-1.01] | p = 0.158 |
| Hemoglobin (per 1 g/dL) | 2769 | 45 | 41 | 0.70 | [0.59-0.83] | p < 0.001 |
| Platelets (per 1000/mm^3^) | 2769 | 45 | 41 | 1.00 | [1.00-1.00] | p = 0.798 |
| White blood cells (per  1000/mm^3^) | 2769 | 45 | 41 | 1.16 | [1.09-1.23] | p < 0.001 |

**Table S3. Breakdown of all endomyocardial biopsy samples by rejection subtype.**

| Rejection subtype | All rejection | Treated rejection |
| --- | --- | --- |
| Acute cellular rejection (ACR) |  |  |
| 0R, samples/total (%) | 1549/2752 (56.3) | 0/1549 (0) |
| 1R, samples/total (%) | 1070/2752 (38.9) | 0/1070 (0) |
| 2R, samples/total (%) | 40/2752 (1.5) | 40/40 (100) |
| 3R, samples/total (%) | 4/2752 (0.1) | 4/4 (100) |
| Antibody mediated rejection (AMR) |  |  |
| pAMR0, samples/total (%) | 2619/2754 (95.1) | 1/2619 (0.04) |
| pAMR1, samples/total (%) | 47/2754 (1.7) | 19/47 (40.4) |
| pAMR2, samples/total (%) | 35/2754 (1.3) | 28/35 (80.0) |
| pAMR3, samples/total (%) | 0/2754 | – |
| Mixed rejection |  |  |
| ACR - 2R/pAMR1, samples/total (%) | 4/2752 (0.1) | 4/4 (100) |
| ACR - 2R/pAMR2, samples/total (%) | 3/2752 (0.1) | 3/3 (100) |

**Table S4. Multi-predictor logistic regression for treated acute rejection for all endomyocardial biopsies.** CI, confidence interval; DSA, donor specific antibody; HTx, heart transplant; OR, odds ratio.

| Variables | OR | 95% CI | p-value |
| --- | --- | --- | --- |
| For cause indication | 9.17 | [4.56-18.46] | p_c_ < 0.001 |
| Concurrent DSA | 5.12 | [2.56-10.25] | p_c_ < 0.001 |
| Inpatient status | 1.74 | [0.93-3.26] | p_c_ = 0.124 |
| Right atrial pressure (per 1 mmHg) | 1.06 | [1.00-1.13] | p_c_ = 0.124 |
| Fick cardiac index (per 1 LPM/m2) | 0.65 | [0.44-0.96] | p_c_ = 0.093 |
| Time since HTx (per week) | 1.00 | [1.00-1.00] | p_c_ = 0.909 |

**Table S5. Breakdown of endomyocardial biopsy samples performed within 1 month of heart transplant.**

| Rejection subtype | Surveillance | Treated surveillance | For cause | Treated for cause |
| --- | --- | --- | --- | --- |
| Acute cellular rejection |  |  |  |  |
| 0R, samples/total (%) | 217/372 (58.3) | – | 25/41 (61.0) | – |
| 1R, samples/total (%) | 151/372 (40.6) | – | 14/41 (34.1) | – |
| 2R, samples/total (%) | 3/372 (0.8) | 3/3 (100) | 2/41 (4.9) | 2/2 (100) |
| 3R, samples/total (%) | 1/372 (0.3) | 1/1 (100) | 0/41 | – |
| Antibody mediated rejection |  |  |  |  |
| pAMR0, samples/total (%) | 368/375 (98.1) | – | 39/48 (81.3) | – |
| pAMR1, samples/total (%) | 7/375 (1.9) | 1/7 (14.3) | 7/48 (14.6) | 6/7 (85.7) |
| pAMR2, samples/total (%) | 0/375 | – | 2/48 (4.2) | 2/2 (100) |
| pAMR3, samples/total (%) | 0/375 | – | 0/48 | – |
| Mixed rejection | 0/375 | – | 0/48 | – |

**Table S6. Causes of death in study patients.**

| Infection |  |
| --- | --- |
| COVID-19 pneumonia, N (%) | 2 (8.3) |
| Bacterial, N (%) | 8 (33.3) |
| Fungal, N (%) | 1 (4.2) |
| Acute rejection |  |
| Acute cellular rejection, N (%) | 0 |
| Antibody mediated rejection, N (%) | 3 (12.5) |
| Cardiac allograft vasculopathy, N (%) | 2 (8.3) |
| Cancer, N (%) | 3 (12.5) |
| Other, N (%) | 5 (20.8) |
| **Total** | 24 |
